# Transient infection with SARS-CoV-2 without induction of systemic immunity

**DOI:** 10.1101/2021.07.07.21260153

**Authors:** Barbara C. Gärtner, Verena Klemis, Tina Schmidt, Martina Sester, Tim Meyer

## Abstract

SARS-CoV-2 testing using PCR is currently used as screening test to guide isolation and contact tracing. Among 1,700 players and staff of the German Bundesliga and Bundesliga 2 who were regularly tested twice weekly, 98 individuals had a positive PCR. Among those, 11 asymptomatic cases were identified who only had a transient single positive PCR of low viral load. As only one out of 11 individuals developed SARS-CoV-2 specific cellular and humoral immunity, this indicates that transient colonization with SARS-CoV-2 may frequently occur without systemic induction of specific adaptive immunity. This knowledge may have implications for management of isolation and contact tracing, which may not be justified in these cases.

## Main text

Detection of SARS-CoV-2 RNA using PCR is the cornerstone diagnostics for acute infection. As this is interpreted as contagiousness, a single positive test is generally followed by isolation and contact tracing to prevent further spread. In May 2020, a special hygiene program was established for elite soccer players and staff after the lockdown to facilitate the restart of the German Bundesliga and Bundesliga 2 with matches behind closed doors.^1,2^ The concept included twice weekly PCR-testing from nasopharyngeal and/or oropharyngeal swabs. Although confirmatory retesting of positive tests was neither mandatory nor encouraged, some teams adopted retesting. Among approximately 1,700 players/staff who were regularly screened, 98 tested positive between 13.09.2020 and 14.01.2021. Among these, 11 asymptomatic cases were identified with an unexpected pattern of a single positive PCR preceded and immediately followed by a series of negative PCRs (table 1). Viral load was rather low (median Ct 36·3 (37·9-28·0). We wondered if this pattern reflected an actual SARS-CoV-2 infection that resulted in the systemic induction of specific humoral and cellular immunity. A median of 42 (21-114) days after the positive PCR, SARS-CoV-2 specific antibodies and CD4 and CD8 T cells were determined as described before.^3^ A systemic immune response was only found in one out of 11 individuals, whereas the others did not mount any specific immunity (Figure 1).

**Figure 1:**
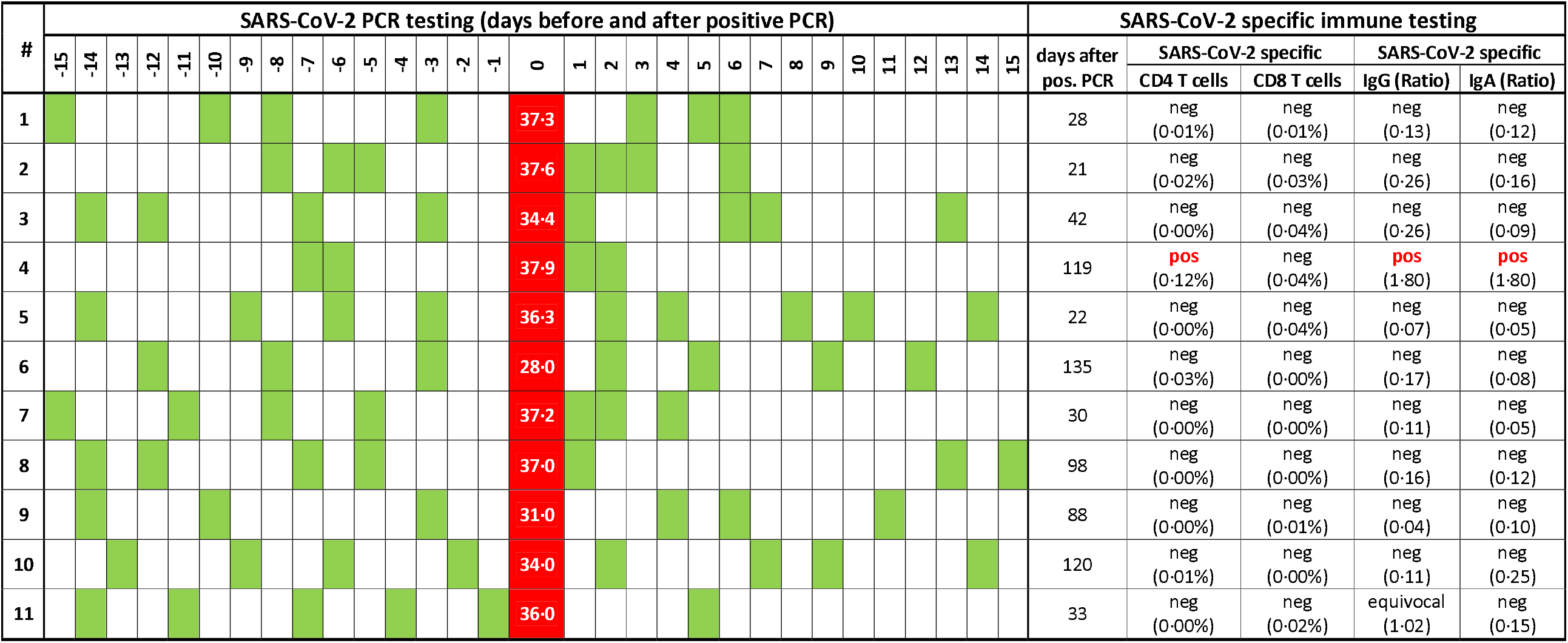
Overview of SARS-CoV-2 PCR test results over time and SARS-CoV-2 specific humoral and cellular immunity. Each line shows data from one individual. All subjects were males aged 30·6±9·1 (mean±standard deviation) years. The chronological order of PCR-testing before and after the positive test is shown with the positive PCR test result including Ct value labeled in red, and negative PCR results in green. SARS-CoV-2 specific IgG and IgA were determined at a median of 42 (range 21-114) days after the positive PCR test using a semiquantitative ELISA (Euroimmun, Lübeck, Germany). Antibody levels are expressed as ratios of the extinction of the sample divided by the extinction of a calibrator serum. Ratios <0·8 were scored negative, ratios between ≥0·8 and <1·1 were scored equivocal, and ratios ≥1·1 were scored positive. Specific CD4 and CD8 T cells were quantified after specific stimulation with overlapping peptide pools as previously described^3^. All individuals showed reactive T cells after stimulation with *Staphylococcus aureus* enterotoxin B, which was used as positive control (CD4 median 2·43% (IQR 1·63-3·62%), CD8 median 6·82% (IQR 4·10-12·94%)). Percentages below 0·05% were scored negative. Individual #4 had negative serological results on screening 3 and 4 months before the positive PCR test, individual #11 had negative serological results on screening 6 months before the positive PCR. Ct, cycle threshold; IQR, interquartile range; PCR, polymerase chain reaction.

A single positive PCR-test in asymptomatic individuals might be a result of a transient colonization confined to the upper respiratory tract.^4^ Local innate immunity may have limited further viral spread and induction of a systemic immune response. Given the absence of symptoms and specific immunity, a false positive PCR test due to contamination during swabbing or testing cannot completely be excluded. However, the high number (at least 11 out of 98 individuals) makes this rather unlikely. Likewise, follow-up samples could have been falsely negative, but this is considered unlikely, as PCR tests remained negative on several occasions.

In conclusion, our unique dataset of regularly screened individuals shows that transient colonization with SARS-CoV-2 may frequently occur in asymptomatic individuals without systemic induction of specific adaptive immunity. This pattern found in more than 10% of positive cases has likely been overlooked so far, since confirmatory retesting shortly after a positive PCR test is generally not recommended and thus rarely done. Further characterization of transient colonization regarding contagiousness is warranted, as preventive measures such as isolation and contact tracing may not be justified in these cases.

## Data Availability

Data will be made available upon request to the corresponding author.

## Acknowledgements

The authors thank all players and staff for their participation, and Candida Guckelmus for expert technical assistance.

## Ethical approval

The study was approved by the ethics committee of the Ärztekammer des Saarlandes (reference 76/20), and all individuals gave written informed consent.

## Authors’ contributions

MS, TM and BG designed that study. VK and TS performed the experiments. All authors analysed the data. MS, TM and BG wrote the manuscript with input of all authors.

